# Preconception *Mycoplasma genitalium* Seropositivity and Risk of Impaired Fecundity

**DOI:** 10.64898/2026.03.03.26347541

**Authors:** Yajnaseni Chakraborti, Stefanie N. Hinkle, Jørgen Skov Jensen, Anna Overgaard Kildemoes, Catherine L. Haggerty, Toni Darville, Sunni L. Mumford, Enrique F Schisterman, Robert M. Silver, John F. Alderete, Brandie DePaoli Taylor

## Abstract

**Background:** *Mycoplasma genitalium (MG)* is an emerging sexually transmitted infection (STI) associated with pelvic inflammatory disease and tubal factor infertility. Its relationship with impaired fecundity remains unclear and is rarely examined in the context of co-seropositivity with other STIs.

**Methods:** We conducted a secondary analysis of the Effects of Aspirin in Gestation and Reproduction (EAGeR) trial, a prospective preconception cohort of women with proven fecundity and prior pregnancy loss. *MG* serostatus was determined using Western blot–based IgG assays on 1146 stored serum specimens. *Chlamydia trachomatis* (CT) and other STIs were also measured. Associations between MG seropositivity and measures of impaired fecundity were assessed.

Pregnancy loss and live birth were modeled using inverse-probability weighted quasi-Poisson and unweighted log-binomial models to calculate relative risks (RR). Fecundability-odds-ratio (FOR) was estimated using a discrete Cox proportional hazards model accounting for left truncation and right censoring. Propensity score (PS) weighted versions of these models assessed risks associated with *CT* co-seropositivity. Analyses were adjusted for demographic and reproductive history variables.

**Results:** Overall, 17.1% (n=210) of participants were *MG* seropositive, with 27.6% (n=58) co-seropositive with *CT*. Compared to STI-seronegative women, *MG* seropositivity was not associated with any outcome, although trends were observed for reduced fecundability (FOR_adj_: 0.87, 95% CI 0.70-1.08) and live birth (RR_adj_: 0.94, 95% CI 0.79-1.11). Co-seropositivity with *CT* was associated with lower likelihood of live birth [Relative Risk (RR)_PS-weighted_: 0.82, 95% CI: 0.70-0.96]. Sensitivity analyses supported the robustness of these findings.

**Conclusions:** Being co-seropositive for *MG* and *CT* preconception may impair fecundity.

## Introduction

Bacterial sexually transmitted infections (STIs) are associated with pelvic inflammatory disease, ectopic pregnancy, infertility, and adverse birth outcomes.^1–5^ Accordingly, routine screening programs for *Chlamydia trachomatis (CT)* and *Neisseria gonorrhoeae (NG)* aim to reduce infertility at the population level.^6,7^ In contrast, less is known about the long-term reproductive burden of *Mycoplasma genitalium (MG).* Therefore, screening is not currently recommended for this often asymptomatic, antibiotic-resistant, and persistent STI.^4,5,8–10^ Data are limited, but seroprevalence studies suggest that 10-20% of the general population has been exposed to *MG,* highlighting its potential as a reproductive health concern.^11–14^

*MG* is strongly associated with pelvic inflammatory disease and moderately associated with tubal factor infertility.^2,15^ Tubal factor infertility results from prior acute genital tract infections that lead to permanent tissue damage and fallopian tube occlusion.^16–18^ Whether prior exposure to *MG* is linked to other indicators of reduced fecundity, such as longer time-to-pregnancy, pregnancy loss, or lower live birth rates remains unclear, as existing studies are rare and report conflicting results.^19,20^ Most studies were conducted in infertility clinics, lacked assessment of early pregnancy loss, and had insufficient adjustment for confounders.^12–14^ In addition, few accounted for exposure to other bacterial STIs, despite evidence that 36-38% of *MG*-infected women are co-infected with *CT*,^21,22^ which may exacerbate upper genital tract sequelae.^23^

Our objective was to assess whether being *MG* seropositive is associated with indicators of impaired fecundity. We also examined whether being co-seropositive for *MG* and *CT* further increases these risks.

## Methods

This study is a secondary analysis of data and serum samples from 1146 participants of the Effects of Aspirin in Gestation and Reproduction (EAGeR) trial (n=1228).^24^ Briefly, the EAGeR study was a multicenter prospective cohort designed to evaluate whether low-dose aspirin improved live birth rates compared with placebo among individuals who had a history of pregnancy loss. Eligible participants were healthy women aged 18 to 40 years who had one or two prior pregnancy losses, had no known infertility diagnosis, and were actively attempting conception. Participants were followed for up to six menstrual cycles while attempting conception or throughout pregnancy if conception occurred. All participants provided informed consent, and the original study was approved by Institutional Review Boards (IRBs) at each participating site. For the present secondary analysis, the University of Texas Medical Branch IRB classified this work as non-human subjects research.

### Exposures

Serum samples collected at the preconception enrollment visit and stored at -80°C were used to determine the presence of *MG*–specific immunoglobulin G (IgG) antibodies by Western blotting. The MG075F1 recombinant protein fragment was used as the antigen and its production, purification, and cut-off determination are described in Kildemoes *et al.* (2025).^25^ This assay, developed to reduce cross-reactivity between *MG* and the closely related ubiquitous respiratory tract pathogen *M. pneumoniae,* has shown an overall specificity of 95.2% and sensitivity of 87.1%.

Serologic testing was also conducted for *CT*, *Trichomonas vaginalis (TV)*, and *NG*. *CT*-specific antibodies were measured by a synthetic peptide ELISA as previously described.^26^ Methods to determine serologic status for *TV* and *NG* can be found in the Supplementary Materials Appendix A. Briefly, a recombinant actin fragment (ACTSOE3) was used to determine the presence of *TV*-specific antibodies in Western blots with an estimated sensitivity of 75% and specificity of 95.2% and for *NG*, purified pili antigen from the F62 strain was applied in a Western blot format.

For the primary analysis, individuals were categorized as *MG* seropositive, *MG* seronegative but seropositive for *CT*, *TV*, and/or *NG*, and seronegative for all STIs measured in this study. As a secondary analysis we categorized *MG* seropositive individuals into mono and co-seropositive groups including: (1) *MG* mono-seropositive, (2) *MG* co-seropositive with *CT*, and (3) *MG* co-seropositive with *TV* and/or *NG*. Additionally, those who were *MG* seronegative but seropositive for *CT*, *TV*, and/or *NG* were further categorized into (4) *CT* mono-seropositive and (5) seropositive for *CT,* and/or *TV,* and/or *NG.* Levels of exposure stratification are illustrated in Figure 1. The group “seronegative for all study STIs” was used as the reference in all analyses.

**Figure 1:**
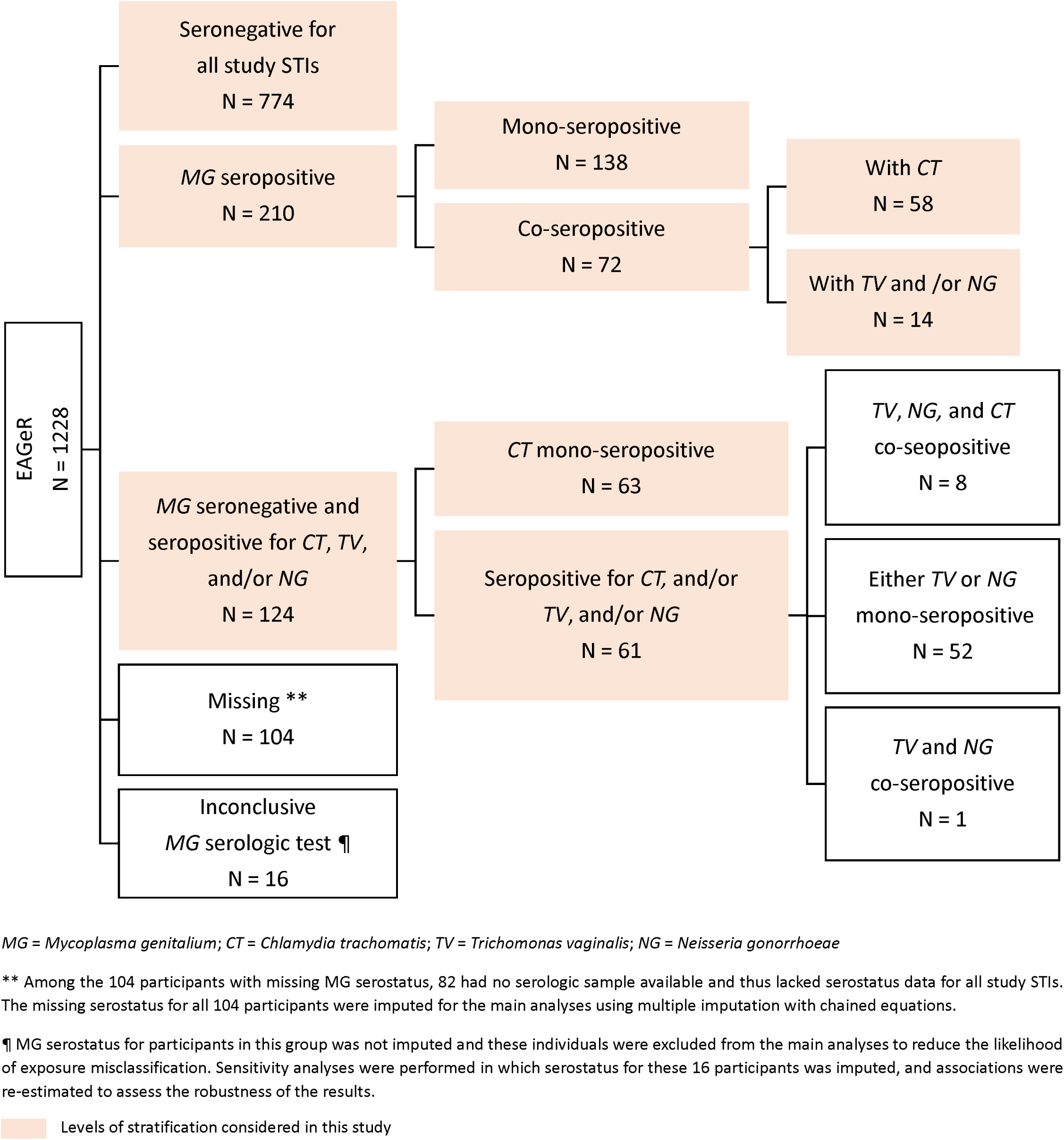
Distribution of serostatus for *M. genitalium, C. trachomatis, T. vaginalis*, and *N. gonorrhoeae*.

### Outcomes

The primary outcomes of interest were live birth and pregnancy loss. Live birth was ascertained through medical record abstraction. Pregnancy loss was defined as either (1) absence of a clinically confirmed pregnancy on ultrasound at 6.5 weeks’ gestation following a positive hCG test (conducted at home and in clinic at the expected menses) or (2) an observed loss after clinical confirmation.

Secondary outcomes included fecundability, hypertensive disorders of pregnancy (HDP), preterm birth, and time-to-delivery (TTD). Fecundability, defined as the probability of pregnancy within one menstrual cycle, was evaluated as the number of menstrual cycles until a positive β-hCG test. HDP, determined from medical records, included preeclampsia or gestational hypertension diagnosed at ≥20 weeks’ gestation, excluding participants with chronic hypertension (n=11). Preterm birth was defined as a live birth occurring between 20 and 36 weeks+6 days of gestation. TTD for live births was estimated using gestational age at delivery, determined by medical records, early pregnancy ultrasound, or menstrual cycle dating with fertility monitors.

At enrollment, participants completed questionnaires on socio-demographic factors, lifestyle behaviors, medical and reproductive history, and family medical history. Based on prior literature, the following were included a priori as potential confounders: age, body mass index (BMI), race, Spanish/Hispanic/Latino ethnicity, marital status, education, annual income, health insurance, smoking status in the past year, alcohol consumption in the past year, and number of sexual partners. Smoking, BMI, and alcohol consumption in the past year, as well as number of sexual partners^27^ were considered proxies for behavioral and lifestyle factors, while race/ethnicity, education, annual income, and health insurance were treated as social determinants of health associated with *MG* exposure.^28–30^

### Statistical Analysis

Baseline participant characteristics and study outcomes were summarized by the primary analysis exposure groups. Outcome distributions were also summarized by *TV* and *NG* serostatus. Outcome distributions by *CT* serostatus have been summarized previously.^26^

Missing data for covariates, serostatus, and outcomes were imputed using multiple imputation with chained equations. Of note, serostatus was not imputed for participants with inconclusive *MG* assay results to reduce the likelihood of exposure misclassification. Logistic regression was used to impute binary variables, and predictive mean matching was applied for continuous and time-to-event variables. Under the Missing at Random assumption, 10 imputed datasets were generated and used to estimate pooled fecundability odds ratios, hazard ratios, and risk ratios.

After excluding participants with inconclusive *MG* serological tests, the association between *MG* seropositivity and fecundability was modeled using discrete Cox proportional hazards models, accounting for cycles before randomization without conception (left truncation) and right censoring. The effect of *MG* seropositivity on live birth was assessed using log-binomial models.

For outcomes conditional on conception or pregnancy progression (pregnancy loss, HDP, preterm birth, and TTD), the cohort was subset accordingly. For example, analyses of pregnancy loss were restricted to participants with β-hCG-confirmed conception. To address potential selection bias introduced by conditioning on conception (β-hCG-positivity), loss-to-follow-up, or pregnancy survival beyond 20 weeks, inverse probability weights (IPW) computed via Generalized Boosted Models^31^ were applied in these analyses.

A weighted quasi-Poisson model was used to estimate the relative risk of pregnancy loss, applying combined IPWs calculated by multiplying the IPWs for β-hCG-positive pregnancies and IPWs for loss-to-follow-up. The relative risks for HDP and preterm birth were estimated using log-binomial models, with IPWs reflecting the combined likelihood of β-hCG positivity, loss-to-follow-up, and pregnancy survival beyond 20 weeks. A discrete-time Cox PH model was used to estimate the hazard of shorter gestational age at delivery, incorporating the same set of IPWs. All IPW and outcome models were adjusted for baseline demographic and reproductive history variables.

The same analytical models were applied when examining secondary exposures, including *MG* mono-seropositivity and co-seropositivity. Because stratification reduced sample sizes, we used propensity score (PS) weighting to balance baseline confounders across exposure groups and preserve exchangeability, rather than adjusting directly in the regression models. The propensity score weights were computed via gradient boosted logistic regression (adjusted for baseline confounders), which accommodates more than two exposure groups. When both IPWs and PS weights were required, combined weights were calculated by multiplying the propensity score weights by the applicable IPWs. Furthermore, to evaluate potential bias from unmeasured variables, we estimated E-values^32^ post-hoc for outcomes showing elevated risks associated with primary or secondary exposures. We also conducted sensitivity analyses wherein *MG* serostatus for participants with inconclusive assay results was imputed, and the models were rerun to assess the robustness of our original findings.

All data cleaning, multiple imputation, and statistical analyses were carried out using R version 4.3.3 (2024-02-29 ucrt), with the boosted models implemented via the R package *twang*.^31^

## Results

In this study of 1228 women actively trying to conceive with proven fecundity and a history of 1-2 pregnancy losses, 63% (n=774) were seronegative for all study STIs, 17.1% (n=210) were *MG* seropositive, and 10.9% (n=124) were seronegative for *MG* but seropositive for *CT*, *TV*, and/or *NG*. Sixteen (1.3%) of the 1228 EAGeR participants had an inconclusive *MG* serological test and were excluded from the analysis to avoid any bias resulting from exposure misclassification. Of the 104 (8.5%) participants with missing *MG* serostatus, 82 had no serologic sample available. Among those seropositive for *MG*, co-seroprevalence with *CT* was 27.6% (n = 58), co-seroprevalence with *TV* was 8.6% (n=18), and co-seroprevalence with *NG* was 6.2% (n=13) (Supplementary Table 1).

Table 1 shows that *MG* seropositive individuals were slightly older, had higher BMI, and were less likely to be married or have more than a high school education, compared to those who were seronegative for all study STIs. Additionally, they were more often uninsured, had lower income levels, reported a greater number of sexual partners, and had used tobacco or consumed alcohol in the past 12 months. Participants who were seronegative for *MG* but seropositive for *CT*, *TV*, and/or *NG* also differed modestly across these characteristics, compared to those seronegative for all study STIs. These differences were similar in direction and magnitude to those observed in *MG* seropositive individuals.

**Table 1.** Sample characteristics with respect to *M. genitalium* serostatus.

| Demographic Characteristics<sup>¶</sup> |  | Serostatus – N (%)<sup>¶</sup> |  |  |  |
| --- | --- | --- | --- | --- | --- |
|  |  | Seronegative for all study STIs | MG seropositive | MG seronegative but seropositive for CT, TV, and/or NG | Missing* |
|  |  | 774 (63.0%) | 210 (17.1%) | 124 (10.9%) | 104 (8.5%) |
| Age (years) | Mean (SD) | 28.8 (4.7) | 29.2 (4.9) | 28.5 (5.3) | 28.4 (4.6) |
|  | Missing – N (%) | - | - | - | - |
| BMI | Median (IQR) | 24.2 (21.5, 28.7) | 26 (21.7, 33) | 25.8 (21.7, 30.1) | 25.2 (22.1, 31.7) |
|  | Missing – N (%) | 10 (1.3%) | - | 3 (2.4%) | 3 (2.9%) |
| Married | Yes | 736 (95.1%) | 171 (81.4%) | 107 (86.3%) | 95 (91.3%) |
|  | No | 38 (4.9%) | 39 (18.6%) | 17 (13.7%) | 9 (8.7%) |
|  | Missing – N (%) | - | - | - | - |
| White | Yes | 749 (96.8%) | 189 (90.0%) | 111 (89.5%) | 97 (93.3%) |
|  | No | 25 (3.2%) | 21 (10.0%) | 13 (10.5%) | 7 (6.7%) |
|  | Missing – N (%) | - | - | - | - |
| Spanish/Hispanic /Latino | Yes | 728 (94.1%) | 186 (88.6%) | 111 (89.5%) | 98 (94.2%) |
|  | No | 46 (5.9%) | 24 (11.4%) | 12 (9.7%) | 6 (5.8%) |
|  | Missing – N (%) | - | - | 1 (0.8%) | - |
| Education | <= High School | 83 (10.7%) | 38 (18.1%) | 28 (22.6%) | 18 (17.3%) |
|  | > High School | 690 (89.1%) | 172 (81.9%) | 96 (77.4%) | 86 (82.7%) |
|  | Missing – N (%) | 1 (0.2%) | - | - | - |
| Income | < \$19,999 | 49 (6.3%) | 25 (11.9%) | 9 (7.3%) | 10 (9.6%) |
| | \$20,000 - \$39,999 | 180 (23.3%) | 63 (30.0%) | 36 (29.0%) | 27 (26.0%) |
| | \$40,000 - \$74,999 | 115 (14.9%) | 38 (18.1%) | 18 (14.5%) | 10 (9.6%) |
| | \$75,000 - \$99,999 | 102 (13.2%) | 19 (9.0%) | 16 (12.9%) | 9 (8.7%) |
| | >= \$100,000 | 327 (42.2%) | 65 (31.0%) | 45 (36.3%) | 48 (46.2%) |
|  | Missing – N (%) | 1 (0.2%) | - | - | - |
| Smoke | Daily/Fewer | 70 (9.0%) | 55 (26.2%) | 17 (13.7%) | 6 (5.8%) |
|  | Never | 697 (90.1%) | 153 (72.9%) | 106 (85.5%) | 97 (93.3%) |
|  | Missing – N (%) | 7 (0.9%) | 2 (1.0%) | 1 (0.8%) | 1 (1.0%) |
| Alcohol Intensity | Never | 525 (67.8%) | 119 (56.7%) | 70 (56.5%) | 78 (75.0%) |
|  | Sometimes/Often | 237 (30.6%) | 90 (42.9%) | 53 (42.7%) | 25 (24.0%) |
|  | Missing – N (%) | 12 (1.6%) | 1 (0.5%) | 1 (0.8%) | 1 (1.0%) |
| Number of sexual partners | Median (IQR) | 1 (1, 4) | 4 (1, 8) | 3 (1, 10) | 1 (1, 5) |
|  | Missing – N (%) | 23 (3.0%) | 13 (6.2%) | 6 (4.8%) | 6 (5.8%) |
| Health Insurance | Yes | 702 (90.7%) | 175 (83.3%) | 102 (82.3%) | 95 (91.3%) |
|  | No | 71 (9.2%) | 34 (16.2%) | 22 (17.7%) | 8 (7.7%) |
|  | Missing – N (%) | 1 (0.1%) | 1 (0.5%) | - | 1 (1.0%) |

MG = *Mycoplasma genitalium*; CT = *Chlamydia trachomatis*; TV = *Trichomonas vaginalis*; NG = *Neisseria gonorrhoeae*
¶ 16 (1.3%) of the 1228 EAGeR participants had inconclusive MG serological test and were excluded from the analysis to decrease likelihood of exposure misclassification.
¶ Percentages within each demographic characteristic category (when applicable) were calculated using the respective serostatus sample size as the denominator.
\* Among the 104 participants with missing MG serostatus, 82 had no serologic sample available and thus lacked serostatus data for all study STIs.

As shown in Table 2, observed β-hCG-detected pregnancy (58.6% vs 68.9%) and live birth rates (41.4% vs 53.6%) were lower in the *MG* seropositive group compared with those seronegative for all study STIs. Median time-to-β-hCG-detected pregnancy was one cycle longer among *MG* seropositive individuals. Rates of pregnancy loss (26.9% vs 21.3%) and preterm birth (10.3% vs 8.7%) were also higher in the *MG* seropositive group. In contrast, HDP (8%) and average gestational age at delivery (38.8 weeks) were the same for both groups. Those seronegative for *MG* but seropositive for *CT*, *TV*, and/or *NG* had pregnancy outcomes largely comparable to those with *MG* seropositivity. However, on closer examination, individuals with *CT* mono-seropositivity showed higher percentages of pregnancy loss and lower percentages of live birth compared to the other exposure groups considered in the secondary analysis (Table 3). Further, individuals co-seropositive for *MG* and *CT* had a higher frequency of adverse pregnancy outcomes compared to those who were *MG* mono-seropositive or *MG* co-seropositive with *TV* and/or *NG*.

**Table 2.** Primary and secondary outcomes with respect to *M. genitalium* serostatus.

| Outcomes of Interest |  | Serostatus – N (%)<sup>¶</sup> |  |  |  |
| --- | --- | --- | --- | --- | --- |
|  |  | Seronegative for all study STIs | MG seropositive | MG seronegative but seropositive for CT, TV, and/or NG | Missing* |
| **Primary Outcomes** |  |  |  |  |  |
| **Live birth** <sup>‡</sup> | Yes | 415 (53.6%) | 87 (41.4%) | 48 (38.7%) | 40 (38.5%) |
|  | No | 284 (36.7%) | 85 (40.5%) | 53 (42.7%) | 53 (51.0%) |
|  | Missing – N (%) | 75 (9.7%) | 38 (18.1%) | 23 (18.5%) | 11 (10.5%) |
| **Any pregnancy loss** <sup>‡‡</sup> | Yes | 112 (21.3%) | 32 (26.9%) | 24 (33.3%) | 19 (32.2%) |
|  | No | 415 (78.7%) | 87 (73.1%) | 48 (66.7%) | 40 (67.8%) |
|  | Missing – N (%) | - | - | - | - |
| **Secondary Outcomes** |  |  |  |  |  |
| **β-hCG detected pregnancy** <sup>‡</sup> | Yes | 533 (68.9%) | 123 (58.6%) | 73 (58.9%) | 60 (57.7%) |
|  | No | 241 (31.1%) | 87 (41.4%) | 51 (41.1%) | 44 (42.3%) |
|  | Missing – N (%) | - | - | - | - |
| **Number of menstrual cycles to β-hCG detected pregnancy** <sup>‡</sup> | Median (IQR) | 5 (3, 8) | 6 (4, 9) | 5 (3, 8) | 6 (4, 10) |
|  | Missing – N (%) | 53 (6.8%) | 17 (8.1%) | 12 (9.7%) | 14 (13.5%) |
| **Hypertensive disorders** ** | Yes | 33 (7.9%) | 7 (8.0%) | 6 (11.8%) | 4 (9.5%) |
|  | No | 380 (90.9%) | 78 (88.6%) | 44 (86.3%) | 36 (85.7%) |
|  | Missing – N (%) | 5 (1.2%) | 3 (3.4%) | 1 (1.9%) | 2 (4.8%) |
| **Preterm birth** <sup>‡</sup> | Yes | 36 (8.7%) | 9 (10.3%) | 5 (10.4%) | 2 (5.0%) |
|  | No | 377 (90.8%) | 78 (89.7%) | 43 (89.6%) | 38 (95.0%) |
|  | Missing – N (%) | 2 (0.5%) | - | - | - |
| **Gestational age at delivery in weeks** <sup>‡</sup> | Mean (SD) | 38.8 (1.8) | 38.8 (2.5) | 38.4 (1.9) | 39.0 (1.2) |
|  | Missing – N (%) | 2 (0.5%) | - | - | - |
| MG = *Mycoplasma genitalium*; CT = *Chlamydia trachomatis*; TV = *Trichomonas vaginalis*; NG = *Neisseria gonorrhoeae*<br>¶ Percentages within each outcome category were calculated using the respective serostatus sample size as the denominator.<br>* Among the 104 participants with missing MG serostatus, 82 had no serologic sample available and thus lacked serostatus data for all study STIs.<br>‡ Sample size N = 1212 (Seronegative = 774; MG seropositive = 210; MG seronegative but seropositive for CT, TV, and/or NG = 124; Missing = 104. 16 (1.3%) of the 1228 EAGeR participants with inconclusive MG serological test were excluded.)<br>‡‡ Sample size N = 777 (Seronegative = 527; MG seropositive = 119; MG seronegative but seropositive for CT, TV, and/or NG = 72; Missing = 59. Among the 797 participants who had a positive pregnancy test, 8 participants with inconclusive MG serological test and 12 participants who were lost-to-follow-up were excluded.)<br>** Sample size N = 599 (Seronegative = 418; MG seropositive = 88; MG seronegative but seropositive for CT, TV, and/or NG = 51; Missing = 42. Among the 797 participants who had a positive pregnancy test, 8 participants with inconclusive MG serological test, 12 participants who were lost-to-follow-up, and 178 whose pregnancy lasted < 20 weeks were excluded.)<br>Hypertensive disorders were defined as presence of preterm/term preeclampsia or gestational hypertension but no evidence of chronic hypertension.<br>‡ Sample size N = 597 (Seronegative = 415; MG seropositive = 87; MG seronegative but seropositive for CT, TV, and/or NG = 48; Missing = 40. Among the 797 participants who had a positive pregnancy test, 8 participants with inconclusive MG serological test, 12 participants who were lost-to-follow-up, 178 whose pregnancy lasted < 20 weeks and 2 with any pregnancy loss were excluded.) |  |  |  |  |  |

**Table 3.**
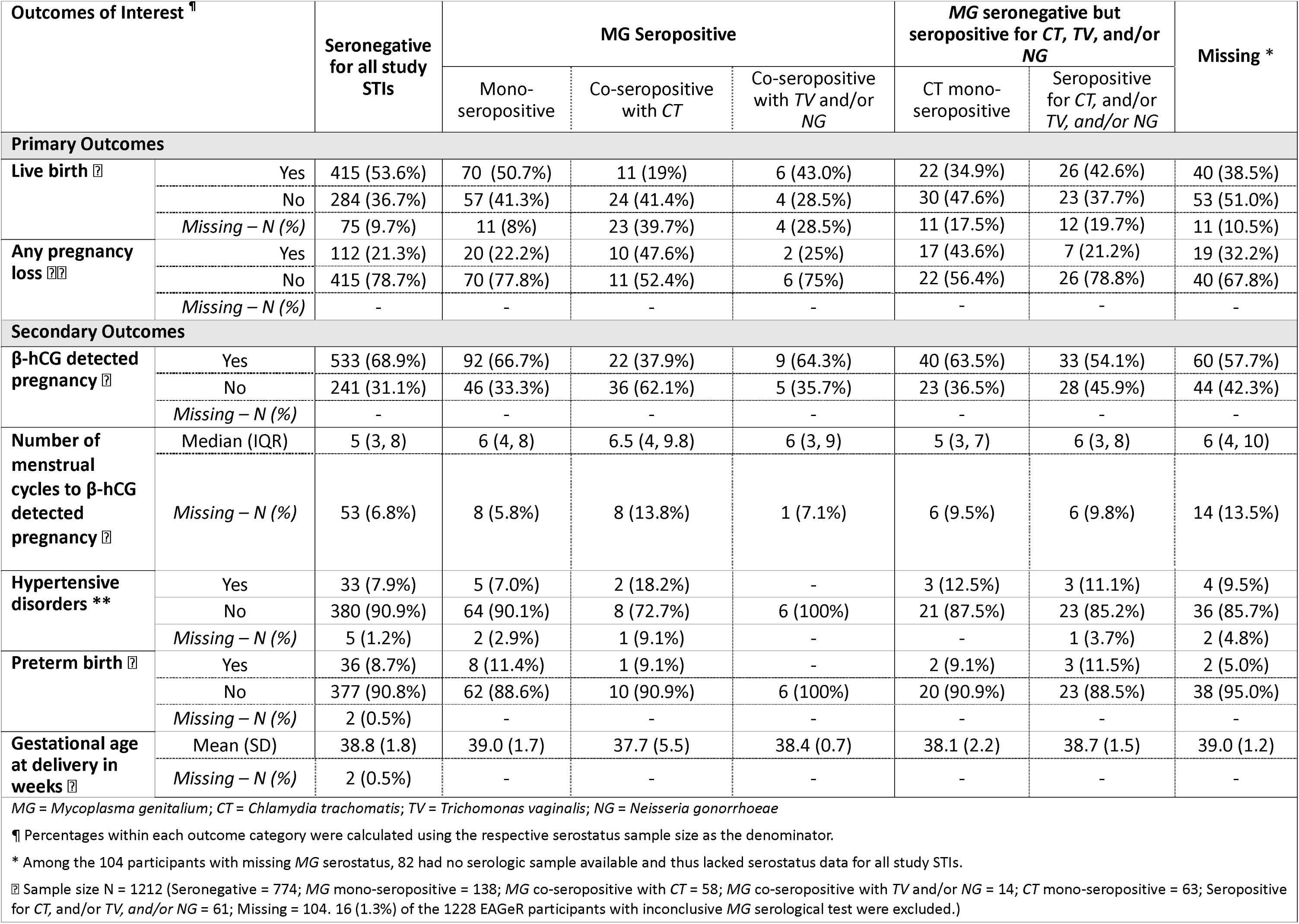
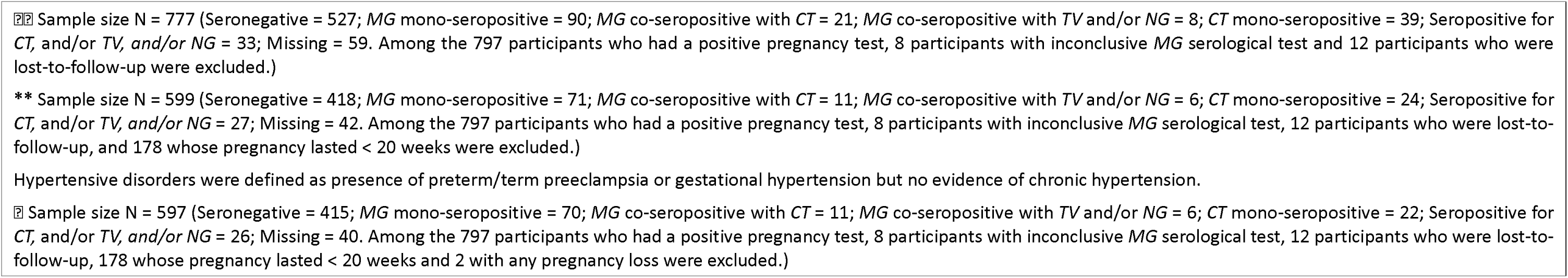
*M. genitalium* mono/co-serostatus with *C. trachomatis* and adverse pregnancy outcomes.

| Outcomes of Interest <sup>¶</sup> |  | Seronegative<br>for all study<br>STIs | MG Seropositive |  |  | MG seronegative but<br>seropositive for CT, TV, and/or<br>NG |  | Missing * |
| --- | --- | --- | --- | --- | --- | --- | --- | --- |
|  |  |  | Mono-seropositive | Co-seropositive<br>with CT | Co-seropositive<br>with TV and/or<br>NG | CT mono-seropositive | Seropositive<br>for CT, and/or<br>TV, and/or NG |  |
| Primary Outcomes |  |  |  |  |  |  |  |  |
| Live birth <sup>‡</sup> | Yes | 415 (53.6%) | 70 (50.7%) | 11 (19%) | 6 (43.0%) | 22 (34.9%) | 26 (42.6%) | 40 (38.5%) |
|  | No | 284 (36.7%) | 57 (41.3%) | 24 (41.4%) | 4 (28.5%) | 30 (47.6%) | 23 (37.7%) | 53 (51.0%) |
|  | Missing – N (%) | 75 (9.7%) | 11 (8%) | 23 (39.7%) | 4 (28.5%) | 11 (17.5%) | 12 (19.7%) | 11 (10.5%) |
| Any pregnancy<br>loss <sup>‡‡</sup> | Yes | 112 (21.3%) | 20 (22.2%) | 10 (47.6%) | 2 (25%) | 17 (43.6%) | 7 (21.2%) | 19 (32.2%) |
|  | No | 415 (78.7%) | 70 (77.8%) | 11 (52.4%) | 6 (75%) | 22 (56.4%) | 26 (78.8%) | 40 (67.8%) |
|  | Missing – N (%) | - | - | - | - | - | - | - |
| Secondary Outcomes |  |  |  |  |  |  |  |  |
| β-hCG detected<br>pregnancy <sup>‡</sup> | Yes | 533 (68.9%) | 92 (66.7%) | 22 (37.9%) | 9 (64.3%) | 40 (63.5%) | 33 (54.1%) | 60 (57.7%) |
|  | No | 241 (31.1%) | 46 (33.3%) | 36 (62.1%) | 5 (35.7%) | 23 (36.5%) | 28 (45.9%) | 44 (42.3%) |
|  | Missing – N (%) | - | - | - | - | - | - | - |
| Number of<br>menstrual<br>cycles to β-hCG<br>detected<br>pregnancy <sup>‡</sup> | Median (IQR) | 5 (3, 8) | 6 (4, 8) | 6.5 (4, 9.8) | 6 (3, 9) | 5 (3, 7) | 6 (3, 8) | 6 (4, 10) |
|  | Missing – N (%) | 53 (6.8%) | 8 (5.8%) | 8 (13.8%) | 1 (7.1%) | 6 (9.5%) | 6 (9.8%) | 14 (13.5%) |
| Hypertensive<br>disorders ** | Yes | 33 (7.9%) | 5 (7.0%) | 2 (18.2%) | - | 3 (12.5%) | 3 (11.1%) | 4 (9.5%) |
|  | No | 380 (90.9%) | 64 (90.1%) | 8 (72.7%) | 6 (100%) | 21 (87.5%) | 23 (85.2%) | 36 (85.7%) |
|  | Missing – N (%) | 5 (1.2%) | 2 (2.9%) | 1 (9.1%) | - | - | 1 (3.7%) | 2 (4.8%) |
| Preterm birth <sup>‡</sup> | Yes | 36 (8.7%) | 8 (11.4%) | 1 (9.1%) | - | 2 (9.1%) | 3 (11.5%) | 2 (5.0%) |
|  | No | 377 (90.8%) | 62 (88.6%) | 10 (90.9%) | 6 (100%) | 20 (90.9%) | 23 (88.5%) | 38 (95.0%) |
|  | Missing – N (%) | 2 (0.5%) | - | - | - | - | - | - |
| Gestational age<br>at delivery in<br>weeks <sup>‡</sup> | Mean (SD) | 38.8 (1.8) | 39.0 (1.7) | 37.7 (5.5) | 38.4 (0.7) | 38.1 (2.2) | 38.7 (1.5) | 39.0 (1.2) |
|  | Missing – N (%) | 2 (0.5%) | - | - | - | - | - | - |

MG = *Mycoplasma genitalium*; CT = *Chlamydia trachomatis*; TV = *Trichomonas vaginalis*; NG = *Neisseria gonorrhoeae*
¶ Percentages within each outcome category were calculated using the respective serostatus sample size as the denominator.
\* Among the 104 participants with missing MG serostatus, 82 had no serologic sample available and thus lacked serostatus data for all study STIs.
‡ Sample size N = 1212 (Seronegative = 774; MG mono-seropositive = 138; MG co-seropositive with CT = 58; MG co-seropositive with TV and/or NG = 14; CT mono-seropositive = 63; Seropositive for CT, and/or TV, and/or NG = 61; Missing = 104. 16 (1.3%) of the 1228 EAGeR participants with inconclusive MG serological test were excluded.)
☒☒ Sample size N = 777 (Seronegative = 527; *MG* mono-seropositive = 90; *MG* co-seropositive with *CT* = 21; *MG* co-seropositive with *TV* and/or *NG* = 8; *CT* mono-seropositive = 39; Seropositive for *CT*, and/or *TV*, and/or *NG* = 33; Missing = 59. Among the 797 participants who had a positive pregnancy test, 8 participants with inconclusive *MG* serological test and 12 participants who were lost-to-follow-up were excluded.)
\*\* Sample size N = 599 (Seronegative = 418; *MG* mono-seropositive = 71; *MG* co-seropositive with *CT* = 11; *MG* co-seropositive with *TV* and/or *NG* = 6; *CT* mono-seropositive = 24; Seropositive for *CT*, and/or *TV*, and/or *NG* = 27; Missing = 42. Among the 797 participants who had a positive pregnancy test, 8 participants with inconclusive *MG* serological test, 12 participants who were lost-to-follow-up, and 178 whose pregnancy lasted < 20 weeks were excluded.)
Hypertensive disorders were defined as presence of preterm/term preeclampsia or gestational hypertension but no evidence of chronic hypertension.
☒ Sample size N = 597 (Seronegative = 415; *MG* mono-seropositive = 70; *MG* co-seropositive with *CT* = 11; *MG* co-seropositive with *TV* and/or *NG* = 6; *CT* mono-seropositive = 22; Seropositive for *CT*, and/or *TV*, and/or *NG* = 26; Missing = 40. Among the 797 participants who had a positive pregnancy test, 8 participants with inconclusive *MG* serological test, 12 participants who were lost-to-follow-up, 178 whose pregnancy lasted < 20 weeks and 2 with any pregnancy loss were excluded.)

*MG* seropositivity was associated with lower fecundability (FOR 0.80, 95% CI 0.65-0.98). However, after adjustments, confidence intervals slightly overlapped the null value (FOR_adj_: 0.87, 0.70-1.08). *MG* seropositivity was not significantly associated with any other primary or secondary outcomes before or after adjustment for confounding (Figure 2).

**Figure 2:**
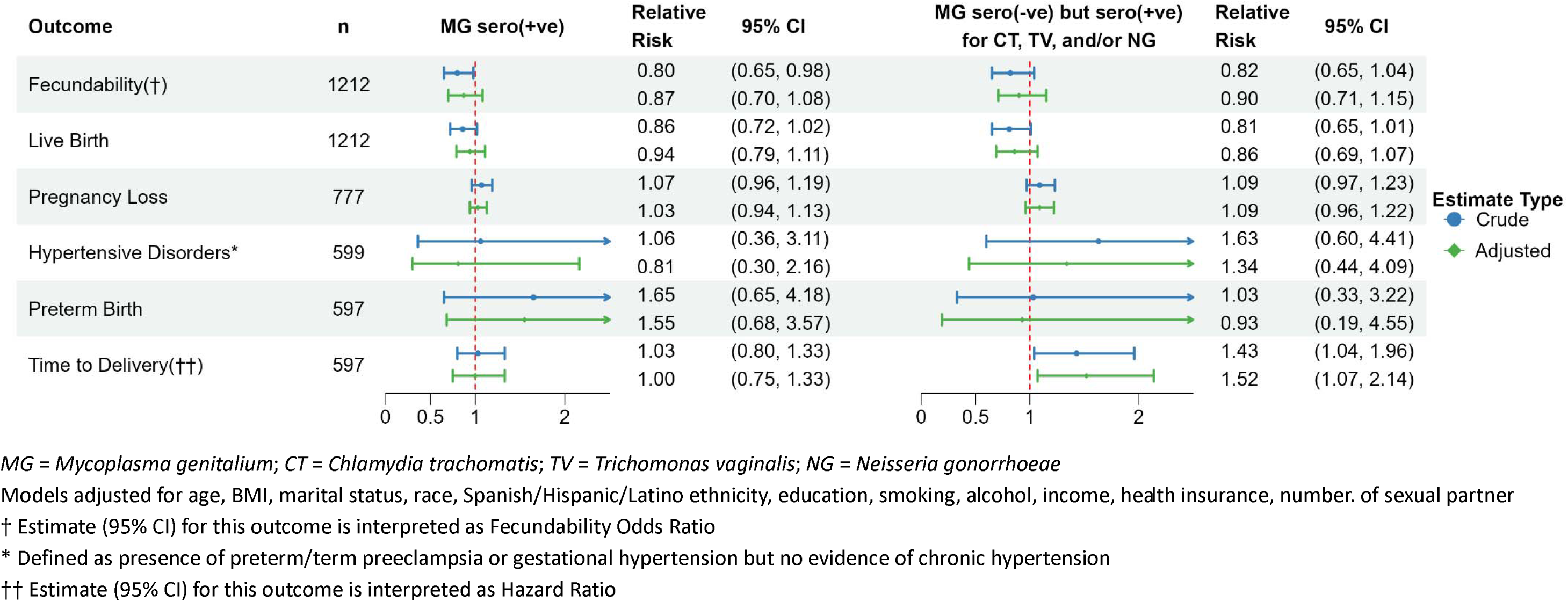
M. genitalium seropositivity and associations with primary and secondary pregnancy outcomes.

Nonetheless, point estimates showed trends toward lower likelihood of live birth (RR_adj_: 0.94, 95% CI: 0.79 – 1.11). Comparable findings were noted among those *MG* seronegative but seropositive for other STIs; this group also showed an association with shorter TTD (HR_adj_: 1.52, 95% CI: 1.07 – 2.14).

Secondary analysis examining the *MG* seropositive group more closely (Table 4) showed that compared with seronegative individuals, those co-seropositive for *MG* and *CT* had a lower likelihood of live birth (RR_PS-weighted_: 0.82, 95% CI: 0.70 – 0.96) and trends toward elevated risk of pregnancy loss (RR_PS-weighted_: 1.20, 95% CI: 0.98 – 1.45) and reduced fecundability (FOR_PS-weighted_: 0.61, 95% CI 0.36-1.03). In contrast, participants co-seropositive for *MG, TV* and/or *NG* appeared to have a lower risk of pregnancy loss (RR_PS-weighted_: 0.84, 95% CI: 0.76 – 0.92) and a higher likelihood of live birth (RR_PS-weighted_: 1.16, 95% CI: 0.99 – 1.35), albeit with borderline statistical significance for the latter. This group also had a trend toward higher fecundability (FOR_PS-weighted_: 1.80, 95% CI: 0.77 – 4.17).

**Table 4:** Primary and secondary pregnancy outcome models stratified by *M. genitalium* mono/co-serostatus in the EAGeR cohort.

| Outcome of interest | Exposure (reference: seronegative for all study STIs) | Crude | Propensity Score Weighted |
| --- | --- | --- | --- |
| **Fecundability Odds Ratio (95% CI)** |  |  |  |
| **Time to 1st HCG detected pregnancy** | *MG* mono-seropositive | 0.88 (0.69 – 1.11) | 0.92 (0.73 – 1.17) |
|  | *MG* co-seropositive with *CT* | 0.52 (0.34 – 0.80) | 0.61 (0.36 – 1.03) |
|  | *MG* co-seropositive with *TV* and/or *NG* | 1.36 (0.70 – 2.67) | 1.80 (0.77 – 4.17) |
|  | *CT* mono-seropositive | 0.77 (0.52 – 1.15) | 0.83 (0.51 – 1.36) |
|  | Seropositive for *CT*, and/or *TV*, and/or *NG* | 0.71 (0.50 – 1.02) | 0.75 (0.52 – 1.07) |
| **Relative Risk (95% CI)** |  |  |  |
| **Live Birth** | *MG* mono-seropositive | 0.93 (0.77 – 1.12) | 0.99 (0.92 – 1.06) |
|  | *MG* co-seropositive with *CT* | 0.54 (0.33 – 0.89) | 0.82 (0.70 – 0.96) |
|  | *MG* co-seropositive with *TV* and/or *NG* | 1.04 (0.63 – 1.72) | 1.16 (0.99 – 1.35) |
|  | *CT* mono-seropositive | 0.83 (0.55 – 1.25) | 0.96 (0.84 – 1.11) |
|  | Seropositive for *CT*, and/or *TV*, and/or *NG* | 0.88 (0.62 – 1.24) | 0.98 (0.86 – 1.12) |
| **Pregnancy Loss <sup>1</sup>** | *MG* mono-seropositive | 1.04 (0.97 – 1.10) | 0.99 (0.87 – 1.12) |
|  | *MG* co-seropositive with *CT* | 1.08 (0.96 – 1.22) | 1.20 (0.98 – 1.45) |
|  | *MG* co-seropositive with *TV* and/or *NG* | 0.98 (0.91 – 1.05) | 0.84 (0.76 – 0.92) |
|  | *CT* mono-seropositive | 1.00 (0.93 – 1.08) | 1.00 (0.77 – 1.32) |
|  | Seropositive for *CT*, and/or *TV*, and/or *NG* | 1.00 (0.94 – 1.07) | 0.97 (0.83 – 1.13) |
| **Hypertensive disorders of pregnancy <sup>1</sup>** | *MG* mono-seropositive | 1.53 (0.36 – 6.40) | 2.15 (0.43 – 10.62) |
|  | *MG* co-seropositive with *CT* | 1.04 (0.20 – 5.51) | 6.06 (0.99 – 37.07) |
|  | *MG* co-seropositive with *TV* and/or *NG* | †† | †† |
|  | *CT* mono-seropositive | 1.40 (0.33 – 5.83) | 1.86 (0.45 – 7.65) |
|  | Seropositive for *CT*, and/or *TV*, and/or *NG* | 1.83 (0.44 – 7.66) | 2.42 (0.48 – 12.21) |
| **Preterm birth <sup>1</sup>** | *MG* mono-seropositive | 2.29 (0.74 – 7.08) | 2.81 (0.76 – 10.44) |
|  | *MG* co-seropositive with *CT* | 2.99 (0.34 – 26.29) | 1.37 (0.15 – 12.37) |
|  | *MG* co-seropositive with *TV* and/or *NG* | †† | †† |
|  | *CT* mono-seropositive | 0.89 (0.16 – 4.99) | 1.27 (0.22 – 7.42) |
|  | Seropositive for *CT*, and/or *TV*, and/or *NG* | 1.15 (0.29 – 4.47) | 1.42 (0.32 – 6.29) |
| **Hazard Ratio (95% CI)** |  |  |  |
| **Time to Delivery (gestational age in weeks) <sup>1</sup>** | *MG* mono-seropositive | 0.93 (0.65 – 1.32) | 0.89 (0.57 – 1.37) |
|  | *MG* co-seropositive with *CT* | 1.17 (0.61 – 2.24) | 1.06 (0.48 – 2.33) |
|  | *MG* co-seropositive with *TV* and/or *NG* | 1.41 (1.00 – 1.99) | 1.41 (1.09 – 1.82) |
|  | *CT* mono-seropositive | 1.24 (0.79 – 1.94) | 1.39 (0.76 – 2.52) |
|  | Seropositive for *CT*, and/or *TV*, and/or *NG* | 1.24 (0.82 – 1.85) | 1.23 (0.83 – 1.82) |
| *MG* = *Mycoplasma genitalium*; *CT* = *Chlamydia trachomatis*; *TV* = *Trichomonas vaginalis*; *NG* = *Neisseria gonorrhoeae*<br><sup>1</sup> Weighted with IPW computed using Generalized Boosted Model<br>†† Not estimable due to inadequate outcome of interest occurring in the exposure group |  |  |  |

Sensitivity analysis indicated that an E-value of 1.74 for the RR_PS-weighted_ point estimate and 1.25 for the confidence interval would be required to nullify the observed association between *MG*-*CT* co-seropositive status and live birth. In other words, an unmeasured confounder would need to be independently associated with both live birth and *MG-CT* co-seropositive status by a risk ratio of at least 1.74 to fully explain away the observed adjusted association. Additionally, to account for even the smallest effect size consistent with our data (i.e., a 4% reduction in live birth likelihood), an unmeasured confounder would need to have a risk ratio of at least 1.25 with both live birth and *MG-CT* co-seropositive status, independently. Further, E-values of 1.67, for the RR_PS-weighted_ point estimate and 1.39 for the confidence interval would be required to nullify the observed association between *MG* co-seropositivity with *TV* and/or *NG* and pregnancy loss, whereas E-values of 1.85 for the HR_PS-weighted_ point estimate and 1.32 for the confidence interval would be required to nullify the observed association with TTD. Finally, sensitivity analyses imputing the serostatus for participants with inconclusive *MG* assay results yielded comparable findings (Supplementary Figure 1 and Supplementary Table 2), further supporting the robustness of results for live birth and pregnancy loss.

We have previously reported results for *CT.*^26^ The number of participants seropositive for *TV* and *NG* was too small for regression analyses. As shown in Supplementary Table 3, among participants seropositive for *TV*, 58.7% (n/N = 37/63) conceived, with a median time-to-β-hCG-detected pregnancy of 5 menstrual cycles; 41.3% (n/N = 26/63) had a live birth, and 27.8% (n/N = 10/37) experienced pregnancy loss. Adverse outcomes were infrequent, including hypertensive disorders of pregnancy (3.7%, n/N = 1/27) and preterm birth (7.7%, n/N = 2/26), with an average gestational age at delivery of 38.7 weeks.

In contrast, outcomes were less favorable among participants seropositive for *NG* (Supplementary Table 4). Only 31.3% (n/N = 10/32) conceived, with a median time-to-β-hCG-detected pregnancy of 6.5 menstrual cycles, the lowest conception rate and longest time to pregnancy observed across all STIs studied. Among this group, 25% (n/N = 8/32) had a live birth, while 20% (n/N = 2/10) experienced pregnancy loss, 25% (n/N = 2/8) developed hypertensive disorders of pregnancy, and 12.5% (n/N = 1/8) had a preterm birth. The average gestational age at delivery was 38.9 weeks.

## Discussion

In this cohort of women with proven fecundity, a history of pregnancy loss, and no known infertility, *MG* seropositivity was not significantly associated with any of the pregnancy outcomes examined. In secondary analyses, participants co-seropositive for *MG* and *CT* had a lower likelihood of live birth, whereas those *MG* co-seropositive with *TV* and/or *NG* had a lower risk of pregnancy loss.

The biological mechanisms linking *MG* to reproductive morbidity are not fully understood. *MG* adheres to the reproductive tract mucosa and can establish chronic infection, leading to tissue damage.^33,34^ It can also induce chronic inflammation^35^ through cycles of persistence and immune activation, potentially causing long-term reproductive tract damage^36^ and increasing susceptibility to other STIs.^37^ Most prior studies have focused on tubal factor infertility,^16–18^ with relatively few evaluating fecundability. For example, a prospective preconception cohort study of 407 Kenyan women reported a 27% lower odds of fecundability among those with NAAT-detected *MG* infection, though results were imprecise (FOR_adj_, 0.73 [95% CI, 0.44, 1.23]).^38^ Similarly, in a study of 461 U.S. women discontinuing contraception, *MG* seropositivity was associated with longer time-to-β-hCG-detected pregnancy (adjusted HR 0.76, 95% CI 0.58–0.99).^39^ Our study also demonstrated a trend toward reduced fecundability, although confidence intervals slightly overlapped the null value after adjustments. Notably, in Peipert et al. (2021),^39^ seropositivity for *MG* was higher at 33%, compared with 17.1% in our study, likely reflecting differences in study populations.

Studies of *MG* and adverse pregnancy outcomes, including pregnancy loss, have primarily focused on active infection. For example, a meta-analysis^19^ reported a significant pooled association between active *MG* infection and pregnancy loss (OR, 1.82 [95% CI, 1.10-3.03]). However, most included studies did not capture early losses and lacked adjustment for key confounders such as co-exposure to other STIs. Although our analytical approach addressed these limitations, we found no evidence of an increased risk of pregnancy loss associated with *MG* alone. A higher risk of pregnancy loss was only indicated among participants co-seropositive for *MG* and *CT*, whereas *MG* co-seropositivity with *TV* and/or *NG* was associated with a lower risk.

In our prior analysis of this cohort, *CT* seropositivity was associated with increased risk of pregnancy loss [RR: 1.16 (95% CI: 1.04, 1.29)] and decreased likelihood of live birth [RR: 0.77 (95% CI: 0.59, 0.99)], but not significantly linked to reduced fecundability [FOR_adj_: 0.92 (95% CI: 0.71, 1.20)].^26^ At the time, measures for other STIs were unavailable, and the reference group included individuals seropositive for other STIs. With the updated data, reduced fecundability and live birth likelihood observed with *MG-CT* co-seropositivity may reflect an amplified effect of both pathogens, as each alone showed weaker but similar trends compared to their combined effect. Such a pattern was less evident for pregnancy loss.

This study has several strengths. First, access to detailed preconception data from a large cohort of women who were actively trying to conceive allowed us to evaluate time-to-pregnancy and capture both early and late pregnancy losses—outcomes not captured in studies of clinically recognized pregnancies. Second, we addressed selection biases that typically arise when subsetting preconception cohorts to examine outcomes conditional on conception. By applying advanced epidemiological weighting methods, we mitigated key methodological limitations common in prior research. Additionally, because stratifying the seropositive participants into mono- and co-seropositive groups reduced sample sizes, we applied propensity score weighting to improve exchangeability across exposure groups. Sensitivity analysis further indicated that only a moderately strong unmeasured confounder could fully account for the observed association between *MG-CT* co-seropositivity and live birth, supporting the robustness of our findings and the appropriate selection of confounders.

The seroprevalence observed in our study was comparable to European populations with a low STI burden.^12–14^ However, most EAGeR participants were recruited in Salt Lake City, Utah, a setting with consistently lower STI rates than the general U.S. population.^40^ While this may limit generalizability, our findings underscore the potential impact of *MG* seropositivity and *CT* co-seropositivity on reproductive outcomes even in a seemingly low-risk population.

In summary, this study evaluated both *MG* seropositivity and co-seropositivity with other STIs in relation to fecundity and other pregnancy outcomes. Our findings suggest that prior exposure to *MG* alone may not lead to adverse reproductive outcomes. However, being *MG-CT* co-seropositive may reduce the likelihood of live birth. By addressing key methodological limitations of previous work, this study provides a framework for future investigations, in larger and more diverse populations. This is particularly important given the high prevalence of antibiotic-resistant *MG* strains, which may lead to persistent infection and long-term reproductive tract damage.

## Data Availability

Data for this study was obtained under a data usage agreement with the Eunice Kennedy Shriver National Institute of Child Health and Human Development and cannot be shared by the authors. Individuals interested in obtaining access to the data can reach out to NICHD to establish a data usage agreement.

## CRediT Authorship Contribution Statement

Yajnaseni Chakraborti (Methodology, Software, Validation, Formal Analysis, Data Curation, Writing – Original/Review & Editing, Visualization); Stefanie N. Hinkle (Methodology, Validation, Resources, Writing – Review & Editing, Supervision); Jørgen Skov Jensen (Conceptualization, Methodology, Investigation, Resources, Writing – Review & Editing); Anna Overgaard Kildemoes (Methodology, Investigation, Writing – Review & Editing); Catherine L. Haggerty (Conceptualization, Investigation, Writing – Review & Editing); Toni Darville (Conceptualization, Investigation, Writing – Review & Editing); Sunni L. Mumford (Methodology, Resources, Data Curation, Writing – Review & Editing); Enrique F Schisterman (Conceptualization, Methodology, Writing – Review & Editing); Robert M. Silver (Data Curation, Writing – Review & Editing); John F. Alderete (Methodology); Brandie DePaoli Taylor (Conceptualization, Methodology, Validation, Investigation, Resources, Data Curation, Writing – Original/Review & Editing, Supervision, Project Administration, Funding Acquisition).

## Attestation Statement

Though information on outcomes from this study population have been published previously, their association with *C. trachomatis* seropositivity has never been published. Data for this study was obtained under a data usage agreement with the Eunice Kennedy Shriver National Institute of Child Health and Human Development and cannot be shared by the authors. Individuals interested in obtaining access to the data can reach out to NICHD to establish a data usage agreement. The STROBE checklist was followed for this study design.

## Data Sharing Statement

Analysis codes in R programming language are available on request.

