## Supplemental Materials for "Preconception *Mycoplasma genitalium* Seropositivity and Risk of Impaired Fecundity"

**Supplementary Materials**

**Appendix A: Measurement of *M. genitalium,* *T. vaginalis* and *N. gonorrhoeae* antibodies**

**Combined detection of IgG specific for *Mycoplasma genitalium* (Mg075F1) and *Trichomonas vaginalis* (ACT-SOE3) antigens by western line blotting**

Cloning, expression, purification, and cut-off determination of the recombinant His-tagged Mg075F1 protein fragment from *M. genitalium*^1^ is described in Kildemoes *et al.* (2025).^2^ The same Mg075F1 batch was used for this study and performed with an overall specificity of 95.2% and sensitivity of 87.1%. For *T. vaginalis*, a recombinant His-tagged α-actinin protein fragment, ACT-SOE3 (Genbank: AAC72899.1, amino acids 259-623; ~72,3 kDa including 6xHis-tag), was used as antigen. Cloning, protein expression and purification of ACT-SOE3 are described in Alderete (2017)^3^. The *T. vaginalis* western lineblot assay was set up and optimized for the detection of IgG anti-ACT-SOE3 based on known negative and positive samples. Estimation of IgG anti-ACT-SOE3 specificity to 95,2% was done based on material from anonymized Danish children (≤15 years old, n=167), who were highly unlikely to have been exposed to *T. vaginalis.* Sensitivity of IgG anti-ACT-SOE3 was estimated to 75,0% based on PCR-confirmed *T. vaginalis-*positive vaginal swab samples (n=32) from Madagascar,^4^ where *T. vaginalis* was detected by real-time PCR as previously described in Pillay *et al.* (2007)^5^ except for use of a modified probe adjusted to a higher T_m_ (FAM-TCATGTCCTCTCCAAGC-MGB). Ethical clearance for use of the Madagascar samples was granted by the Committee of Ethics at the Ministry of Health in Antananarivo (Comité d’Ethique de la Recherche Bio-Médicale auprès du Ministère de la Santé Publique); (Authorization Number: 098-MSANP/CERBM; Number: 059-MSANP/CERBM; Number: 065 MSANP/SG/-AGMED/CNPV/CERBM). Antigen concentrations were estimated by QUBiT protein assay (Invitrogen #Q32857, Thermo Fisher Scientific, Hvidovre, Denmark), and a mix of 1 µg Mg075F1 and 2 µg ACT-SOE3 in 1x Laemmli (Bio-Rad #1610737, Østerbro, Denmark), 50mM DTT, 8M urea was separated on 4-20% TGX gels (precast Mini-Protean TGX, 7cm single well, Bio-Rad #4561091) at 100V constant voltage for 50 min (Bio-Rad #1658004) in standard 25mM Tris, 192mM glycine, 0,1% SDS running buffer (Bio-Rad #1610772). Transfer of the antigen to 0,2 µM nitrocellulose membranes (Bio-Rad #1704158) was done at 25V for three min/gel in a Trans-Blot Turbo Transfer System (Bio-Rad #1704150). The blots were blocked for 30-60 minutes at room temperature and then overnight at 4⁰C in Tris-buffered saline with Tween20 (TBST, Medicago #097510-100, Uppsala, Sweden) with 5% skimmed milk powder (Oxoid #LP0033B, Thermo Fisher Scientific) (blocking/sample buffer). All sera (n=1146) were diluted 1:200 in sample buffer, and 600 µl was loaded onto blots using a Mini-Protean II Multiscreen lineblotter (Bio-Rad #1704017) with capacity for 20 samples per blot and incubated on a shaker for one hour. 41 samples had too high background and were retested in a 1:300 dilution. Aliquots of a constructed pool of known *M. genitalium* and *T. vaginalis* positive sera were used as positive controls on all blots. Blots were washed thrice in TBST for five minutes before incubation with secondary goat-anti-human IgG (y-chain specific) alkaline phosphatase (AP)-conjugated (Sigma-Aldrich #A3188, batch SLBM3258V; 1:5000, Merck Life Science A/S, Søborg, Denmark) in sample buffer on a shaker for 1 hour. Monoclonal mouse IgG2b-anti human 6x-His(c-term) antibody (3D5)-AP (Invitrogen #R932-25, 1:2500, Thermo Fisher Scientific) was used to confirm the location of the recombinant proteins. After three washes for five minutes on a shaker in TBST, development was done with 10 ml of 1-step BCIP/NBT substrate (Thermo Scientific #34042). The reaction was stopped with milli-Q washes, and blots were dried before imaging. If not specified, incubation was done at room temperature. White light images were produced on the G:BOX Chemi-XRQ system (Syngene, Cambridge, United Kingdom) and processed using the GeneTools software (version 4.3.17.0). Data normalization and analysis was done as described in Kildemoes *et al.* (2025) for both antigens^2^ with the *T. vaginalis* image analysis criteria set to a combination cut-off of relative signal normalized to the internal standardized positive control (≥30) combined with mean pixel unit (≥1400) for positive signal. For both MG075F1 and ACT-SOE3, serology status was recorded as positive (ordinal scale categories from 1-4), under detection limit/negative, inconclusive and/or no data. The latter category applied to samples with persistent high background or other technical issues, which made blot-readouts completely uninterpretable.

**Detection of IgG specific for *Neisseria gonorrhoeae* pili antigen by western line blotting**

The *N. gonorrhoeae* F62 (T_1_,T_2_) strain stored in liquid N_2_ was inoculated onto GC Kellogg agar-plates (SSI Diagnostica A/S #31925, Hillerød, Denmark) on day one and incubated overnight at 36,5°C with 5% CO_2_. Day two colonies with T_1_ and T_2_ morphology^6^ as determined by microscopy were picked, replated onto GC Kellogg agar-plates and incubated 36,5°C with 5% CO_2_ for up to a maximum of 24 hours. Day 3; after confirming correct T_1_ and T_2_ morphology of the colonies, all gonococci were harvested and split into three suspensions in 11ml sterile isotonic saline solution. From each suspension, 100 GC Kellogg agar-plates were inoculated with 100 µl, as well as a set of control plates (one GC Kellogg agar-plate for confirmation of morphology and two chocolate agar plates (SSI Diagnostica A/S #700) without antibiotics for control of contamination). Plates were incubated at 36,5°C with 5% CO_2_ for up to a maximum of 24 hours. Additionally, a Gram stain was performed on each suspension. Day four, the sets of 50 GC Kellogg agar-plates were harvested into 20 ml cold 0,16M ethanolamine pH 10,5 (Sigma-Aldrich #8008490100) and kept on ice to avoid autolysis of the cells. The suspensions were whirly-mixed thoroughly for exactly five minutes. Six chocolate agar plates without antibiotics were inoculated to assess potential contamination. The plates were incubated overnight at 36,5°C with 5% CO_2_ for up to a maximum of 24 hours. The remaining suspensions were transferred to sterile 50 ml centrifuge tubes and spun at 12000 x *g* for ten minutes at 4°C. Supernatants were then transferred to sterile ultra-centrifuge tubes and spun for one hour at 50000 g at 4°C. Supernatants were transferred into sterile Erlenmeyer flasks and mixed with equal volumes of 20% saturated ammonium sulphate (Sigma-Aldrich #A4915). The flasks were then placed overnight at 4°C, where the pili antigen crystallized. Day five, the suspensions were transferred from the Erlenmeyer flasks to two sterile Corex glass centrifuge tubes and spun for one hour at 10,000 x *g*. The supernatants were discarded and the sediment resuspended in 3 ml 0,01M Tris pH 7,0 (Invitrogen #AM9850G) with 0,01M sodium azide. The resulting F62 pili antigen was diluted 2000x in PBS pH6,4, sonicated on ice for one minute with an amplitude of 22 microns (MSE sonicator, MSE Supplies, Tucson, AZ, USA) and then stored at 4⁰C. The antigen is storable for several years. The above method is adapted from Hermodson *et al.* (1978).^7^

Antigen concentration was estimated by Qubit protein assay, and 7 µg pili antigen was used per gel. SDS-PAGE and immunoblotting were carried out as described for MG075F1 and ACT-SOE3 with the exception of the secondary detection antibody being polyclonal goat-anti-human IgG (y-chain specific)-AP (Sigma-Aldrich #A3150, batch SLCN2542; 1:4000). A known aliquoted positive control was included on all blots. Presence of IgG anti-pili bands on dried blots was scored and data entered by two independent investigators in a blinded manner by comparing the blots to a reference blot incubated with known samples. Bands were scored as 0 (under detection limit/negative), 1 (very weak band), 2 (low positive), and 3 (positive), and in case of discrepancy in scoring, a third investigator was consulted.

This assay was performed with an estimated specificity of 100% (n=66 Danish children ≤15 years old) and sensitivity of 80% (n=15 known PCR positive samples from the Madagascan sample set described above). Two of the samples used for sensitivity assessment had very low copy numbers, which could indicate inoculation more than infection, and potentially a pre-seroconversion context; hence, the sensitivity may in reality be closer to 92%.

1.	Lind K, Benzon MW, Jensen JS, Clyde WA. A seroepidemiological study of Mycoplasma pneumoniae infections in Denmark over the 50-year period 1946-1995. *Eur J Epidemiol*. 1997;13(5):581-586. doi:10.1023/a:1007353121693

2.	Kildemoes AMO, Rai OSS, Westermann EPL, et al. Detection of human IgG antibodies against Mycoplasma genitalium using a recombinant MG075 antigen. *J Clin Microbiol*. 63(5):e01876-24. doi:10.1128/jcm.01876-24

**Appendix B: Results**

**Supplementary Table 1. Distribution N(%) of *M. genitalium* co-serostatus with *C. trachomatis*, *T. vaginalis*, and *N. gonorrhoeae***

| Other STI seropositivity status^¶^ | | Seronegative for all study STIs | *MG* Seropositivity * | | Missing** |
| --- | --- | --- | --- | --- | --- |
|  |  |  | ***MG* seropositive** | ***MG* seronegative but seropositive for *CT*, *TV*, and/or *NG*** |  |
|  |  | 774 (63.9%) | 210 (17.3%) | 124 (10.2%) | 104 (8.6%) |
| *CT* | Yes | - | 58 (27.6%) | 71 (57.3%) | 4 (3.8%) |
|  | No | 774 (100%) | 152 (72.4%) | 53 (42.7%) | 17 (16.3%) |
|  | *Missing – N (%)* | *-* | *-* | *-* | *83 (79.9%)* |
| *TV* | Yes | - | 18 (8.6%) | 45 (36.3%) | - |
|  | No | 774 (100%) | 192 (91.4%) | 79 (63.7%) | 2 (1.9%) |
|  | *Missing – N (%)* | *-* | *-* | *-* | *102 (98.1%)* |
| *NG* | Yes | - | 13 (6.2%) | 18 (14.5%) | 1 (0.9%) |
|  | No | 774 (100%) | 197 (93.8%) | 106 (85.5%) | 20 (19.2%) |
|  | *Missing – N (%)* | *-* | *-* | *-* | *83 (79.9%)* |
| *MG* = *Mycoplasma genitalium*; *CT* = *Chlamydia trachomatis*; *TV* = *Trichomonas vaginalis*; *NG* = *Neisseria gonorrhoeae*  * 16 (1.3%) of the 1228 EAGeR participants had inconclusive MG serological test and were excluded from the analysis to decrease likelihood of exposure misclassification.  ¶ Percentages within each serostatus category for *C. trachomatis*, *T. vaginalis*, and *N. gonorrhoeae* were calculated using the respective *M. genitalium* serostatus sample size as the denominator.  ** Among the 104 participants with missing *M. genitalium* serostatus, 82 had no serologic sample available and thus lacked serostatus data for all study STIs. | | | | | |

**Supplementary Figure 1: Sensitivity analysis results for primary and secondary outcomes by serostatus in the full EAGeR sample (including imputed serostatus for 16 participants with inconclusive MG serologic tests)**

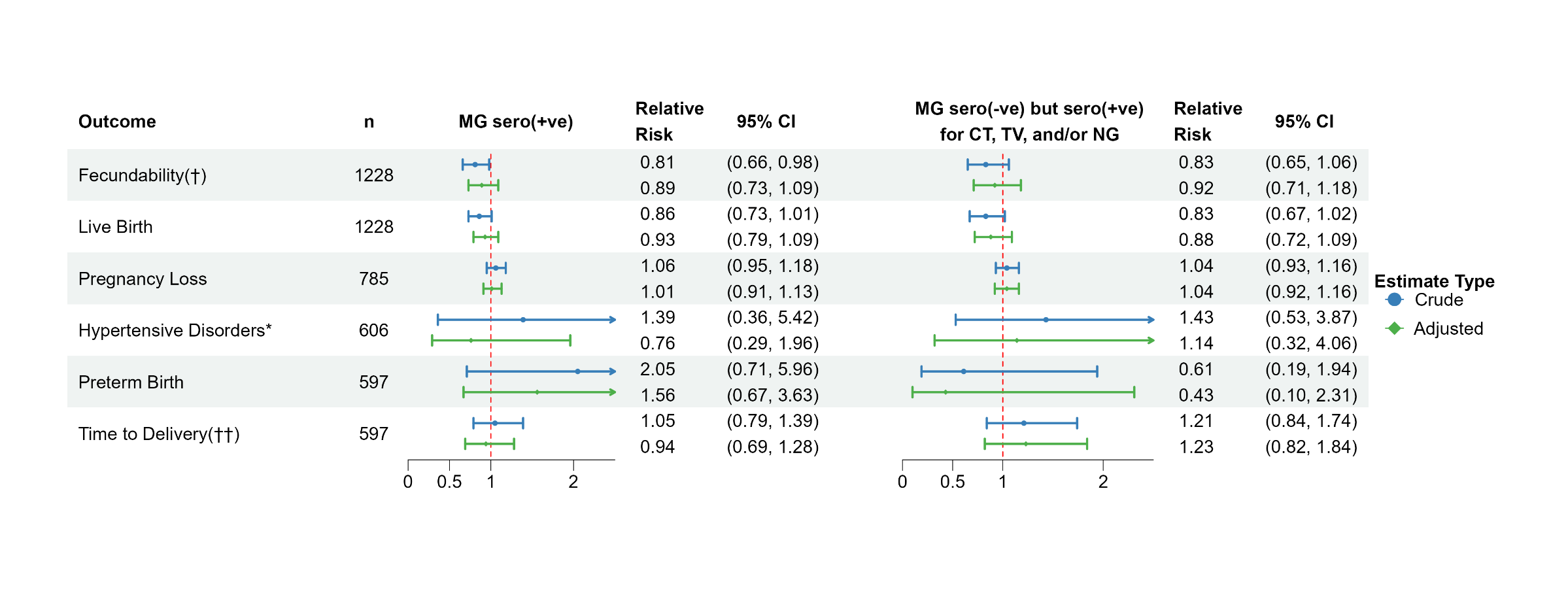

*MG* = *Mycoplasma genitalium*; *CT* = *Chlamydia trachomatis*; *TV* = *Trichomonas vaginalis*; *NG* = *Neisseria gonorrhoeae*

$\dagger$ Estimate (95% CI) for this outcome is interpreted as Fecundability Odds Ratio

* Defined as presence of preterm/term preeclampsia or gestational hypertension but no evidence of chronic hypertension

Models adjusted for age, BMI, marital status, race, Spanish/Hispanic/Latino ethnicity, education, smoking, alcohol, income, health insurance, number. of sexual partner

$\dagger\dagger$ Estimate (95% CI) for this outcome is interpreted as Hazard Ratio

**Supplementary Table 2: Sensitivity analysis—primary and secondary pregnancy outcome models stratified by *M. genitalium* mono/co‑serostatus in the full EAGeR sample (including imputed serostatus for 16 participants with inconclusive MG serologic tests)**

| **Outcome of interest** | **Exposure (reference: seronegative for all STIs)** | **Crude** | **Propensity Score Weighted** |
| --- | --- | --- | --- |
| **Fecundability Odds Ratio (95% CI)** | | | |
| **Time to 1st HCG detected pregnancy** | *MG* mono-seropositive | 0.90 (0.73 – 1.13) | 0.92 (0.73 – 1.15) |
|  | *MG* co-seropositive with *CT* | 0.54 (0.36 – 0.83) | 0.52 (0.28 – 1.00) |
|  | *MG* co-seropositive with *TV* and/or *NG* | 1.10 (0.49 – 2.48) | 0.83 (0.30 – 2.29) |
|  | *CT* mono-seropositive | 0.92 (0.66 – 1.26) | 0.92 (0.52 – 1.63) |
|  | Seropositive for *CT,* and/or *TV, and/or NG* | 0.74 (0.51 – 1.07) | 0.76 (0.49 – 1.19) |
| **Relative Risk (95% CI)** | | | |
| **Live Birth** | *MG* mono-seropositive | 0.93 (0.78 – 1.10) | 0.99 (0.93 – 1.06) |
|  | *MG* co-seropositive with *CT* | 0.58 (0.36 – 0.93) | 0.81 (0.69 – 0.96) |
|  | *MG* co-seropositive with *TV* and/or *NG* | 1.03 (0.56 – 1.89) | 1.03 (0.78 – 1.37) |
|  | *CT* mono-seropositive | 0.73 (0.53 – 1.00) | 0.92 (0.78 – 1.08) |
|  | Seropositive for *CT,* and/or *TV, and/or NG* | 0.94 (0.71 – 1.23) | 0.98 (0.88 – 1.10) |
| **Pregnancy Loss ^1^** | *MG* mono-seropositive | 1.04 (0.92 – 1.16) | 1.00 (0.89 – 1.12) |
|  | *MG* co-seropositive with *CT* | 1.18 (0.98 – 1.43) | 1.22 (0.98 – 1.51) |
|  | *MG* co-seropositive with *TV* and/or *NG* | 0.86 (0.73 – 1.02) | 0.82 (0.75 – 0.89) |
|  | *CT* mono-seropositive | 1.11 (0.96 – 1.28) | 1.06 (0.89 – 1.27) |
|  | Seropositive for *CT,* and/or *TV, and/or NG* | 0.97 (0.82 – 1.15) | 0.95 (0.79 – 1.14) |
| **Hypertensive disorders of pregnancy ^1^** | *MG* mono-seropositive | 1.47 (0.36 – 5.98) | 1.44 (0.36 – 5.79) |
|  | *MG* co-seropositive with *CT* | 0.98 (0.19 – 4.97) | 0.97 (0.19 – 4.89) |
|  | *MG* co-seropositive with *TV* and/or *NG* | $\dagger\dagger$ | $\dagger\dagger$ |
|  | *CT* mono-seropositive | 1.11 (0.29 – 4.21) | 1.11 (0.29 – 4.21) |
|  | Seropositive for *CT,* and/or *TV, and/or NG* | 1.75 (0.47 – 6.51) | 1.73 (0.46 – 6.45) |
| **Preterm birth ^1^** | *MG* mono-seropositive | 2.04 (0.69 – 6.07) | 2.01 (0.68 – 5.92) |
|  | *MG* co-seropositive with *CT* | 2.45 (0.31 – 19.60) | 2.46 (0.31 – 19.70) |
|  | *MG* co-seropositive with *TV* and/or *NG* | $\dagger\dagger$ | $\dagger\dagger$ |
|  | *CT* mono-seropositive | 0.46 (0.10 – 2.14) | 0.47 (0.10 – 2.15) |
|  | Seropositive for *CT,* and/or *TV, and/or NG* | 0.81 (0.16 – 3.97) | 0.80 (0.16 – 3.94) |
| **Hazard Ratio (95% CI)** | | | |
| **Time to Delivery (gestational age in weeks) ^1^** | *MG* mono-seropositive | 0.94 (0.67 – 1.31) | 0.85 (0.55 – 1.32) |
|  | *MG* co-seropositive with *CT* | 1.22 (0.67 – 2.20) | 0.82 (0.41 – 1.65) |
|  | *MG* co-seropositive with *TV* and/or *NG* | 1.38 (0.97 – 1.96) | 1.33 (0.97 – 1.83) |
|  | *CT* mono-seropositive | 1.27 (0.76 – 2.14) | 1.35 (0.72 – 2.53) |
|  | Seropositive for *CT,* and/or *TV, and/or NG* | 1.21 (0.76 – 1.93) | 1.13 (0.73 – 1.73) |
| *MG* = *Mycoplasma genitalium*; *CT* = *Chlamydia trachomatis*; *TV* = *Trichomonas vaginalis*; *NG* = *Neisseria gonorrhoeae*  1 Weighted with IPW computed using Generalized Boosted Model  $\dagger\dagger$ Not estimable due to 0 outcome of interest occurring in the exposure group | | | |

**Supplementary Table 3. Primary and secondary outcomes with respect to *T. vaginalis* seropositivity (n/N = 63/1212)**

| Outcomes of Interest ^¶^ | | | *TV* seropositive |
| --- | --- | --- | --- |
| Primary Outcomes | | | |
| Live birth ꝉ | Yes | 26 (41.3%) | |
|  | No | 20 (31.7%) | |
|  | *Missing – N (%)* | 17 (27.0%) | |
| Any pregnancy loss ꝉꝉ | Yes | 10 (27.8%) | |
|  | No | 26 (72.2%) | |
|  | *Missing – N (%)* | - | |
| Secondary Outcomes | | | |
| β-hCG detected pregnancy ꝉ | Yes | 37 (58.7%) | |
|  | No | 26 (41.3%) | |
|  | *Missing – N (%)* | - | |
| Number of menstrual cycles to β-hCG detected pregnancy ꝉ | Median (IQR) | 5 (3, 8.5) | |
|  | *Missing – N (%)* | 8 (12.7%) | |
| Hypertensive disorders ** | Yes | 1 (3.7%) | |
|  | No | 25 (92.6%) | |
|  | *Missing – N (%)* | 1 (3.7%) | |
| Preterm birth ǂ | Yes | 2 (7.7%) | |
|  | No | 24 (92.3%) | |
|  | *Missing – N (%)* | - | |
| Gestational age at delivery in weeks ǂ | Mean (SD) | 38.7 (1.47) | |
|  | *Missing – N (%)* | - | |
| *TV* = *Trichomonas vaginalis* ¶ Percentages within each category of the primary and secondary outcomes were calculated using the respective sample size (explained below) as the denominator.  ꝉ Sample size N = 63  ꝉꝉ Sample size N = 36 (Among the 63 *T. vaginalis* seropositive EAGeR participants, 37 had a positive pregnancy test and 1 was lost-to-follow-up who was excluded)  ** Sample size N = 27 (Among the 63 *T. vaginalis* seropositive EAGeR participants, 37 had a positive pregnancy test. Among these 37 participants, 1 who was lost-to-follow-up and 9 whose pregnancy lasted < 20 weeks were excluded). Hypertensive disorders were defined as presence of preterm/term preeclampsia or gestational hypertension but no evidence of chronic hypertension.  ǂ Sample size N = 26 (Among the 63 *T. vaginalis* seropositive EAGeR participants, 37 had a positive pregnancy test. Among these 37 participants, 1 who was lost-to-follow-up, 9 whose pregnancy lasted < 20 weeks and 1 with any pregnancy loss were excluded). | | | |

**Supplementary Table 4. Primary and secondary outcomes with respect to *N. gonorrhoeae* seropositivity (n/N = 32/1212)**

| Outcomes of Interest ^¶^ | | | *NG* seropositive |
| --- | --- | --- | --- |
| Primary Outcomes | | | |
| Live birth ꝉ | Yes | 8 (25%) | |
|  | No | 12 (37.5%) | |
|  | *Missing – N (%)* | 12 (37.5%) | |
| Any pregnancy loss ꝉꝉ | Yes | 2 (20%) | |
|  | No | 8 (80%) | |
|  | *Missing – N (%)* | - | |
| Secondary Outcomes | | | |
| β-hCG detected pregnancy ꝉ | Yes | 10 (31.3%) | |
|  | No | 22 (68.7%) | |
|  | *Missing – N (%)* | - | |
| Number of menstrual cycles to β-hCG detected pregnancy ꝉ | Median (IQR) | 6.5 (4, 10.8) | |
|  | *Missing – N (%)* | 2 (6.3%) | |
| Hypertensive disorders ** | Yes | 2 (25%) | |
|  | No | 5 (62.5%) | |
|  | *Missing – N (%)* | 1 (12.5%) | |
| Preterm birth ǂ | Yes | 1 (12.5%) | |
|  | No | 7 (87.5%) | |
|  | *Missing – N (%)* | - | |
| Gestational age at delivery in weeks ǂ | Mean (SD) | 38.9 (1.45) | |
|  | *Missing – N (%)* | - | |
| *NG* = *Neisseria gonorrhoeae*  ¶ Percentages within each category of the primary and secondary outcomes were calculated using the respective sample size (explained below) as the denominator.  ꝉ Sample size N = 32  ꝉꝉ Sample size N = 10 (Among the 32 *N. gonorrhoeae* seropositive EAGeR participants, 10 had a positive pregnancy test).  ** Sample size N = 8 (Among the 32 *N. gonorrhoeae* seropositive EAGeR participants, 10 had a positive pregnancy test. Among these 10 participants, 2 whose pregnancy lasted < 20 weeks were excluded). Hypertensive disorders were defined as presence of preterm/term preeclampsia or gestational hypertension but no evidence of chronic hypertension.  ǂ Sample size N = 8 (Among the 32 *N. gonorrhoeae* seropositive EAGeR participants, 10 had a positive pregnancy test. Among these 10 participants, 2 whose pregnancy lasted < 20 weeks were excluded). | | | |
